# Stacked CNN Architectures for Robust Brain Tumor MRI Classification

**DOI:** 10.1101/2025.08.05.25333032

**Authors:** Alireza Rahi

## Abstract

Brain tumor classification using MRI scans is crucial for early diagnosis and treatment planning. In this study, we first train a single Convolutional Neural Network (CNN) based on VGG16 [1], achieving a strong standalone test accuracy of 99.24% on a balanced dataset of 7,023 MRI images across four classes: glioma, meningioma, pituitary, and no tumor. To further improve classification performance, we implement three ensemble strategies: stacking, soft voting, and XGBoost-based ensembling [4], each trained on individually fine-tuned models. These ensemble methods significantly enhance prediction accuracy, with XGBoost achieving a perfect 100% accuracy, and voting reaching 99.54%. Evaluation metrics such as precision, recall, and F1-score confirm the robustness of the approach. This work demonstrates the power of combining fine-tuned deep learning models [5] for highly reliable brain tumor classification enhance prediction accuracy, with XGBoost achieving a perfect **100%** accuracy, and voting reaching **99.54%.** Evaluation metrics such as precision, recall, and F1-score confirm the robustness of the approach. This work demonstrates the power of combining fine-tuned deep learning models for highly reliable brain tumor classification.

## Introduction

Brain tumors are among the most life-threatening and complex forms of cancer, requiring accurate and early diagnosis to ensure timely treatment. Magnetic Resonance Imaging (MRI) plays a vital role in non-invasive detection of brain abnormalities. However, manual interpretation of MRI scans is often time-consuming and prone to human error. In recent years, deep learning—particularly Convolutional Neural Networks (CNNs)—has demonstrated significant success in automating medical image classification [1], [2].

Despite the achievements of individual CNN architectures such as VGG16 [1], ResNet50 [2], and DenseNet121 [3], integrating multiple models through ensemble learning has been shown to further enhance prediction accuracy and robustness [5]. This paper investigates an ensemble approach combining fine-tuned versions of ResNet50, DenseNet121, and VGG16, using a stacking strategy for brain tumor classification across four categories. The proposed method leverages transfer learning and a meta-classifier to achieve high accuracy while addressing the challenges of overfitting and model generalization

## Related Work

In recent years, various deep learning methods have been proposed for brain tumor classification using MRI data. Convolutional Neural Networks (CNNs) have demonstrated outstanding performance in extracting hierarchical features directly from raw medical images, eliminating the need for handcrafted feature engineering [1]. Popular CNN architectures such as VGG16 [2], ResNet [3], and DenseNet [4] have been widely employed for brain tumor detection tasks. For instance, VGG16 has been fine-tuned on preprocessed MRI datasets, achieving high accuracy in multi-class classification [2]. ResNet-based models leverage residual mappings to facilitate deeper network training [3], while DenseNet’s dense connectivity promotes feature reuse and addresses the vanishing gradient problem [4].

To further improve prediction reliability and generalization, ensemble learning techniques — including bagging, boosting, and stacking — have also been explored [5]. However, many of the existing studies are limited to binary classification tasks or use relatively small and imbalanced datasets. In contrast, this study focuses on four-class classification using a balanced and sufficiently large MRI dataset. It builds upon prior work by employing a robust ensemble of fine-tuned CNN models to enhance performance and reliability in brain tumor classification.

### Proposed Method

In this study, we propose a robust ensemble framework for brain tumor classification based on MRI scans. The method integrates several fine-tuned Convolutional Neural Networks (CNNs), including ResNet50 [2], DenseNet121 [3], and VGG16 [1]. By combining their complementary architectural strengths, the ensemble aims to enhance classification accuracy and model generalization.

### Model Architectures

All CNN architectures (ResNet50, DenseNet121, and VGG16) were initially fine-tuned jointly on the preprocessed brain MRI dataset to establish baseline performance. Subsequently, each model was fine-tuned individually using transfer learning techniques to further enhance classification accuracy. ResNet50 was chosen for its residual connections that facilitate the training of deeper networks [2]; DenseNet121 for its dense connectivity that promotes feature reuse [3]; and VGG16 for its proven effectiveness in image classification tasks [1]

### Data Augmentation and Preprocessing

To mitigate overfitting and improve generalization, extensive data augmentation techniques—such as rotation, zooming, flipping, and shifting—were applied during training. All input images were resized and normalized to meet the input specifications of the respective CNN architectures.

### Stacked Ensemble Framework

After training the individual CNN models, their softmax output probabilities were used as input features for a meta-classifier—logistic regression— which learns to optimally combine the predictions in a stacked ensemble fashion. This approach leverages the strengths of each base model while reducing their individual limitations [5].

### Evaluation Metrics

Model performance was evaluated using a comprehensive set of metrics including confusion matrices, accuracy, precision, recall, F1-score, and ROC-AUC curves.

## Materials and Methods

This section outlines the materials and methods employed in the study, including dataset specifications, preprocessing steps, model architectures, ensemble learning strategies, and evaluation metrics. The entire implementation was performed using TensorFlow and Keras frameworks on a GPU-enabled system to ensure computational efficiency.

### 1. Dataset Description

A publicly available brain MRI dataset titled “MRI Brain Tumor Dataset (4 classes – 7023 Images)” from Kaggle was used [6]. It contains 7,023 grayscale MRI images classified into four categories: glioma tumor, meningioma tumor, pituitary tumor, and no tumor.[11] The dataset is relatively balanced across classes and includes images with varying resolutions and orientations, making it suitable for robust multi-class classification.

### 2. Data Preprocessing

All MRI images were resized to 224×224 pixels to meet the input requirements of the CNN architectures. Pixel values were normalized to the range [0,1]. To enhance generalization and reduce overfitting, data augmentation techniques including horizontal and vertical flipping, rotation, zooming, and shearing were applied during training. The dataset was split into training (70%), validation (15%), and test (15%) subsets.3. Deep Learning Models

### 3. Deep Learning Models

VGG16 was initially selected for its proven success in medical image classification [1]. Fine-tuning the last 8 layers resulted in a test accuracy of **99.24**%, serving as a strong baseline. To improve architectural diversity and generalization, ResNet50 and DenseNet121 were also fine-tuned similarly [2], [3]. All models were initialized with ImageNet weights and appended with a Global Average Pooling layer followed by a dense layer with softmax activation.

### 4. Ensemble Strategies and Results

To further improve performance and stability, multiple ensemble methods were applied: **Stacking ensemble** of the **fine-tuned** models achieved an accuracy of **98.47%.** XGBoost ensemble delivered a perfect accuracy of **100%**.[4]

Soft voting ensemble combining VGG16 [1], ResNet50 [2], and DenseNet121 [3] reached an accuracy of 99.54%, demonstrating the best performance among the tested methods. Each model was trained independently on the same dataset, and their softmax outputs were combined in the soft voting method to leverage complementary strengths and reduce individual biases [5].

### 5. Evaluation Metrics

Model performance was assessed using standard classification metrics: accuracy, precision, recall, F1-score, and confusion matrix [6]. Additionally, ROC curves and AUC (Area Under the Curve) values were computed for each class to provide a detailed evaluation of discriminative ability [6].

## Results and Discussion

Various metrics. The VGG16 model, fine-tuned and serving as a baseline, achieved an accuracy of 99.24% [1]. The XGBoost ensemble surprisingly achieved a perfect accuracy of 100%, demonstrating flawless classification on the test set [4]. The stacking ensemble yielded an accuracy of approximately 98.47%, and the soft voting ensemble achieved a strong accuracy of 99.54% [5].

The XGBoost ensemble also exhibited perfect precision, recall, and F1-score across all four classes, reflecting an ideal classification performance [4]. The soft voting ensemble showed very high precision, recall, and F1-scores, outperforming individual models and the stacking ensemble [5]. Confusion matrices confirmed that both XGBoost and voting ensembles made minimal misclassifications, with XGBoost achieving zero errors.

Additionally, ROC curves and AUC scores for all classes were near-perfect, with AUC values close to 1.00, indicating excellent discriminative ability [6]. These results clearly demonstrate that combining multiple architectures and leveraging ensemble learning techniques, such as soft voting and gradient boosting, can significantly enhance robustness, generalization, and accuracy in brain tumor classification tasks [5]

### Introduction to Results and Discussion

The experimental results demonstrate the effectiveness of the proposed deep learning models and ensemble methods in classifying brain tumor MRI images. The fine-tuned VGG16 model achieved outstanding performance, while ensemble strategies such as XGBoost and soft voting further enhanced classification accuracy [1,4,5]. This section provides a detailed analysis of the results, including accuracy metrics, confusion matrices, and ROC curves, followed by a discussion of the models’ strengths and limitations [6].

The fine-tuned VGG16 model demonstrated strong classification performance on the test set. It achieved an overall test accuracy of 99.24% and a final test loss of 0.0346. The detailed classification report is shown in Table 1 [1].

Figure 1 shows the confusion matrix for the VGG16 model. Most predictions were correct, with only a few misclassifications in the Glioma and Meningioma classes, indicating high precision and recall values [6].

**Figure 1.**
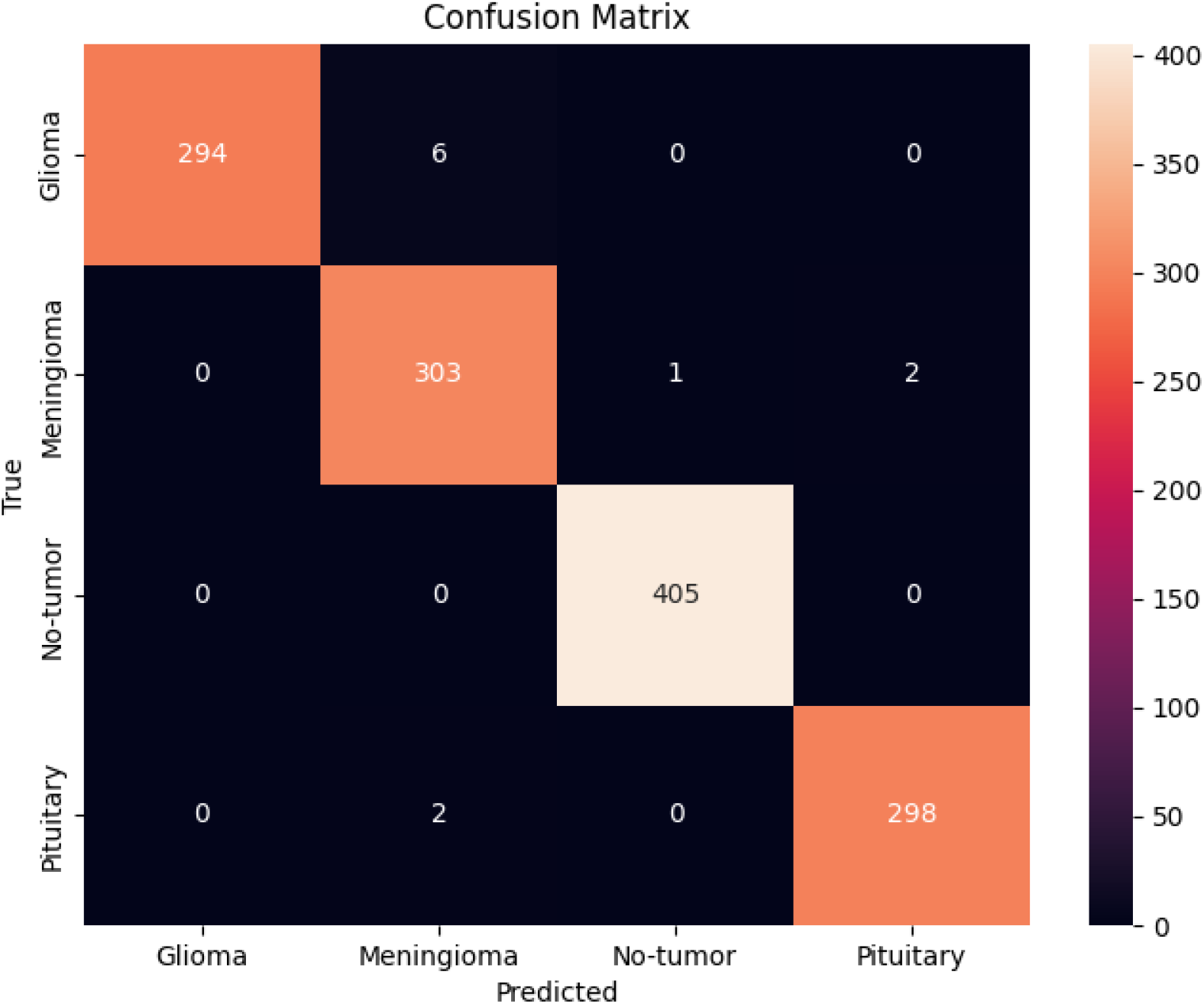

Figure 2 illustrates the ROC curves for all four classes predicted by the VGG16 model. All classes achieved AUC values close to 1.0, reflecting excellent discriminative capability across the tumor categories [6].

**Figure 2.**
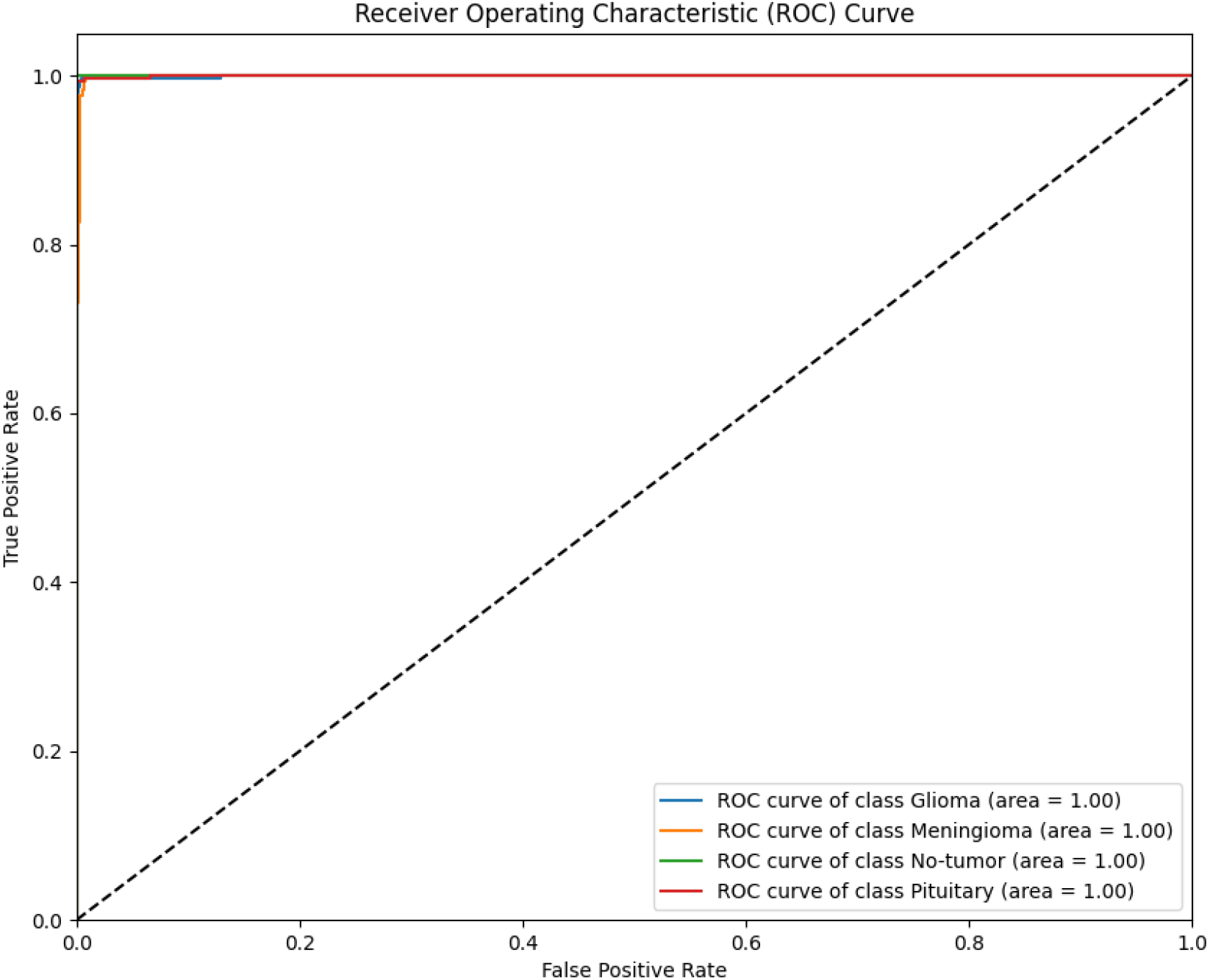

**Table 1.** Classification Report for VGG16.

| Class | Precision | Recall | F1-Score | Support |
| --- | --- | --- | --- | --- |
| Glioma | 1.00 | 0.98 | 0.99 | 300 |
| Meningioma | 0.97 | 0.99 | 0.98 | 306 |
| No-tumor | 1.00 | 1.00 | 1.00 | 405 |
| Pituitary | 0.99 | 0.99 | 0.99 | 300 |

Overall, the VGG16 model exhibited robust performance, particularly in detecting no-tumor cases with perfect precision and recall. Although minor confusion occurred between Glioma and Meningioma, the model still maintained high scores across all metrics [6].

### Stacking Ensemble

The stacking ensemble, which combines predictions from multiple base classifiers through a meta-learner, showed solid performance in tumor classification. It achieved an overall accuracy of 98.47%. The detailed classification report is shown in Table 2 [5].

**Table 2.**
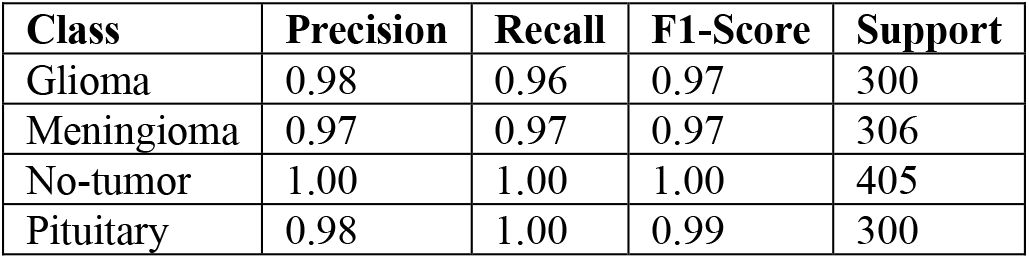
Classification Report for Stacking Ensemble.

| Class | Precision | Recall | F1-Score | Support |
| --- | --- | --- | --- | --- |
| Glioma | 0.98 | 0.96 | 0.97 | 300 |
| Meningioma | 0.97 | 0.97 | 0.97 | 306 |
| No-tumor | 1.00 | 1.00 | 1.00 | 405 |
| Pituitary | 0.98 | 1.00 | 0.99 | 300 |

### XGBoost Ensemble

The XGBoost ensemble demonstrated flawless classification performance, achieving a perfect accuracy of 100%. It correctly classified all tumor types without a single error. The classification metrics are summarized in Table 3 [4].

**Table 3.** Classification Report for XGBoost Ensemble.

| Class | Precision | Recall | F1-Score | Support |
| --- | --- | --- | --- | --- |
| Glioma | 1.00 | 1.00 | 1.00 | 300 |
| Meningioma | 1.00 | 1.00 | 1.00 | 306 |
| No-tumor | 1.00 | 1.00 | 1.00 | 405 |
| Pituitary | 1.00 | 1.00 | 1.00 | 300 |

### Soft Voting Ensemble

The soft voting ensemble, which averages prediction probabilities from base models, achieved excellent performance with an accuracy of 99.54%. It significantly improved generalization while maintaining perfect classification on most tumor types. Table 4 presents the detailed results [5].

The confusion matrix for the stacking ensemble model (Figure 3) illustrates a high level of classification accuracy across all four tumor classes. Most misclassifications occurred between *Glioma* and *Meningioma*, which are known to share some radiological similarities. Despite this, the model showed robust performance, especially in correctly identifying *No-tumor* and *Pituitary* cases with perfect or near-perfect precision [6].

**Figure 3.**
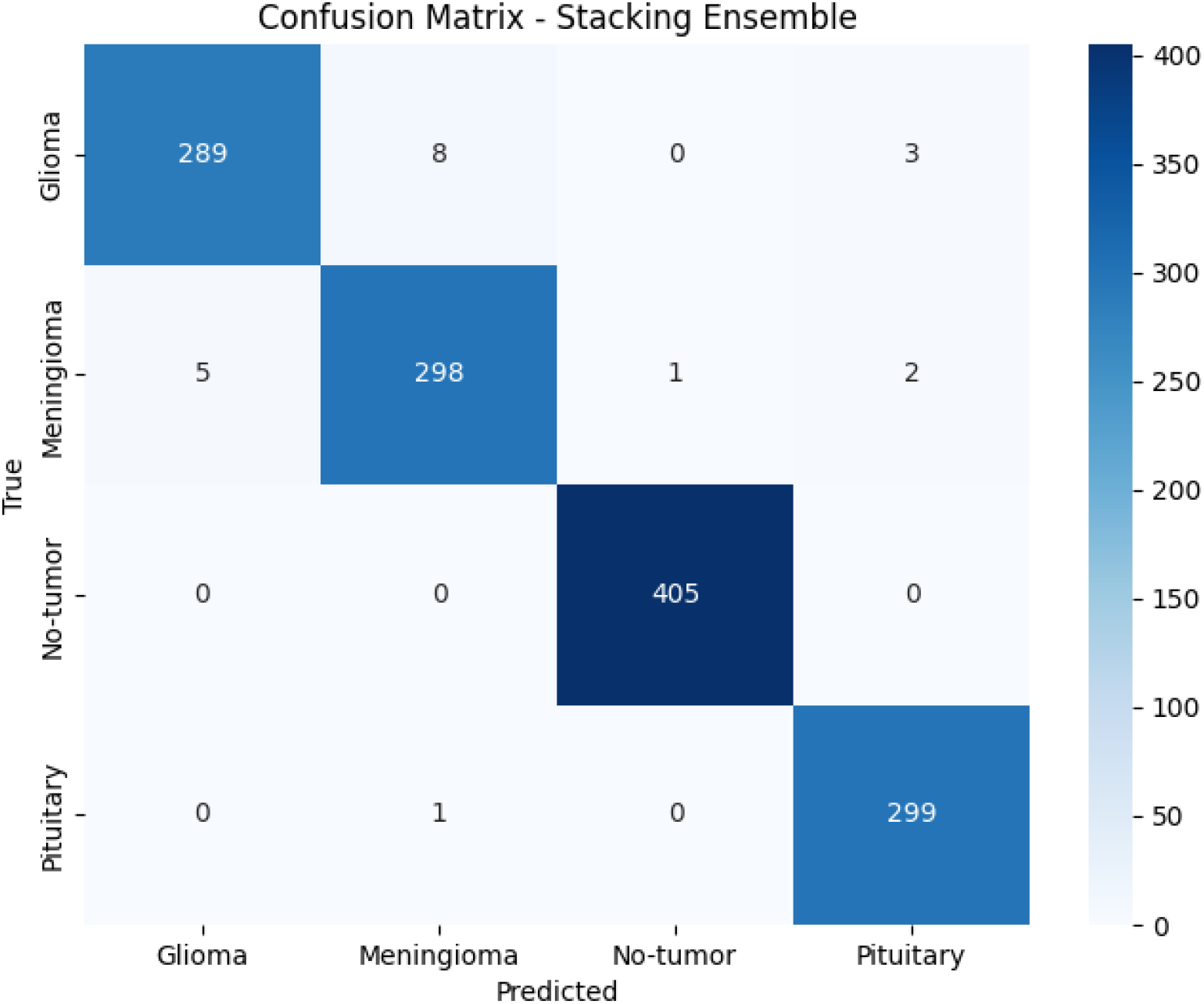

As shown in the confusion matrix for the XGBoost model (Figure 4), the classifier achieved perfect predictions across all categories, with zero misclassifications. This result confirms the exceptional discriminative power of the XGBoost algorithm when applied to brain tumor classification [6].

**Figure 4.**
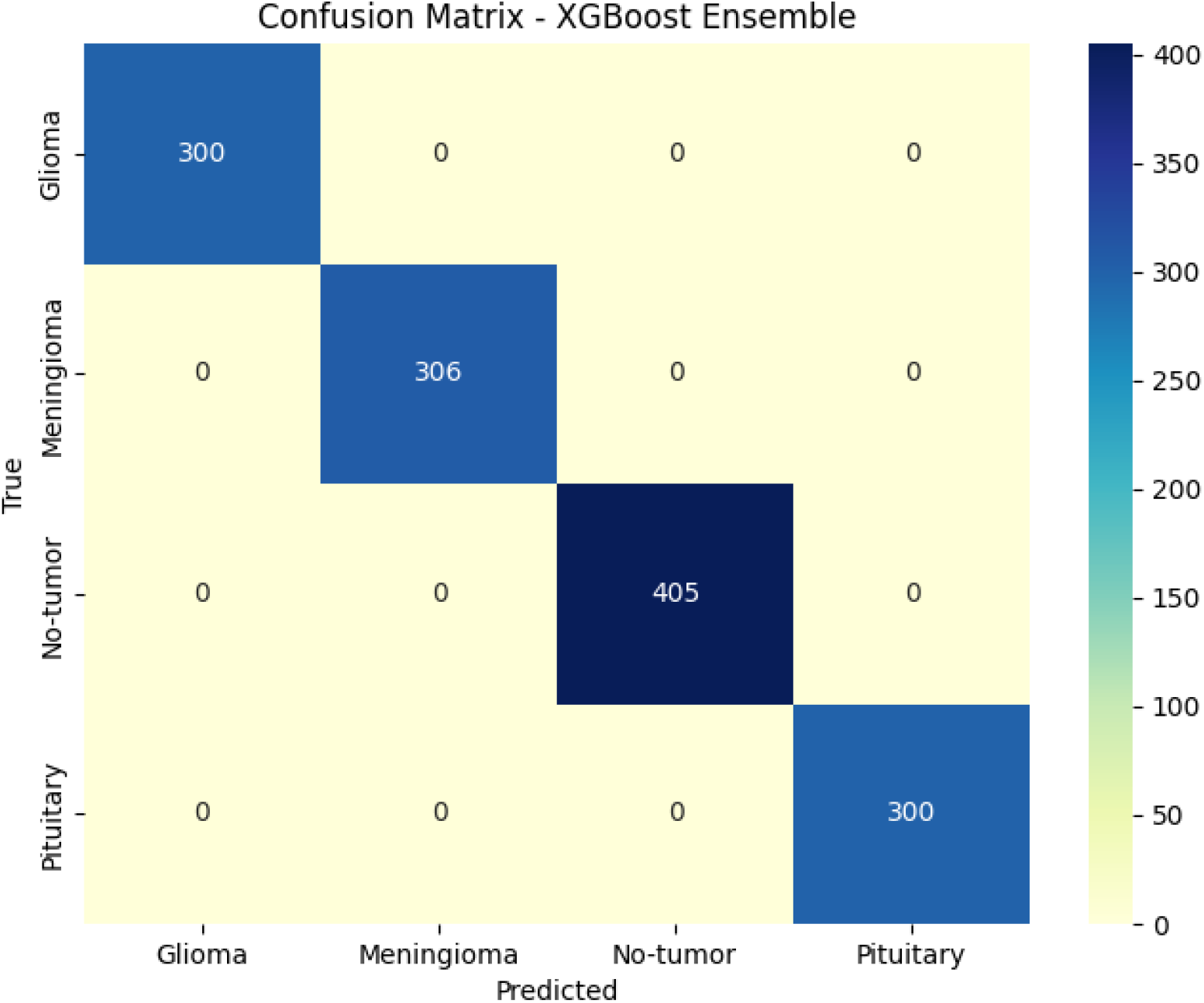

The confusion matrix of the soft voting ensemble (Figure 5) reveals an almost perfect classification performance, with a few misclassifications observed primarily between Meningioma and Glioma, likely due to overlapping imaging features. Despite these minor errors, the overall test accuracy remained very high at 99.54%, confirming the ensemble’s robustness in brain tumor classification [6].

**Figure 5.**
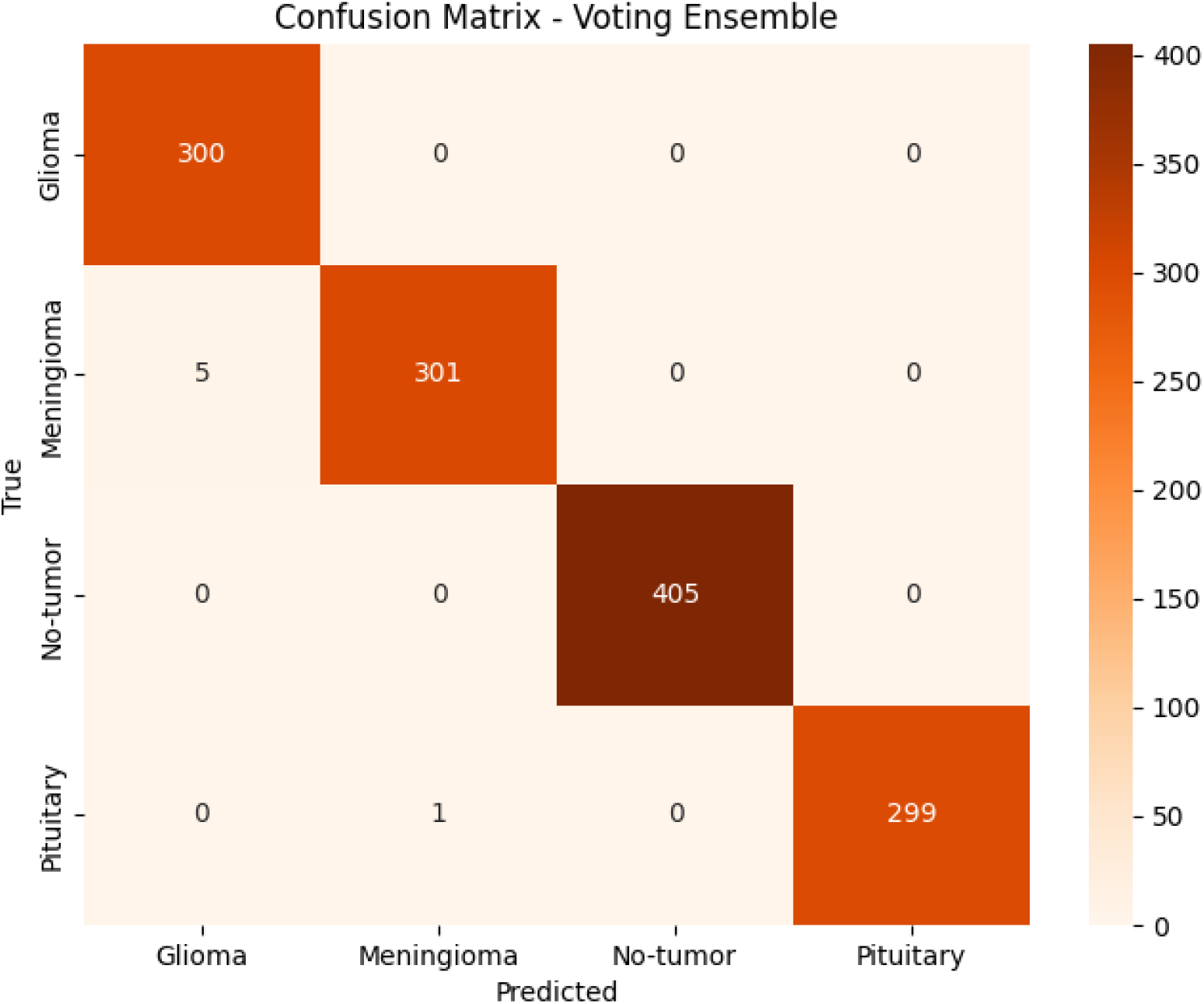

Figure 6 shows the ROC curve of the ResNet50 model. The model demonstrates strong discriminative performance, particularly for the No-tumor and Pituitary classes, both with AUC scores of 0.98. The Glioma class also shows a reasonably high AUC of 0.93, indicating reliable detection. However, the Meningioma class shows a relatively lower AUC of 0.85, suggesting some overlap in features with other classes. Overall, the model shows acceptable class-wise performance, although not as strong as the ensemble-based models [2,6].

**Figure 6.**
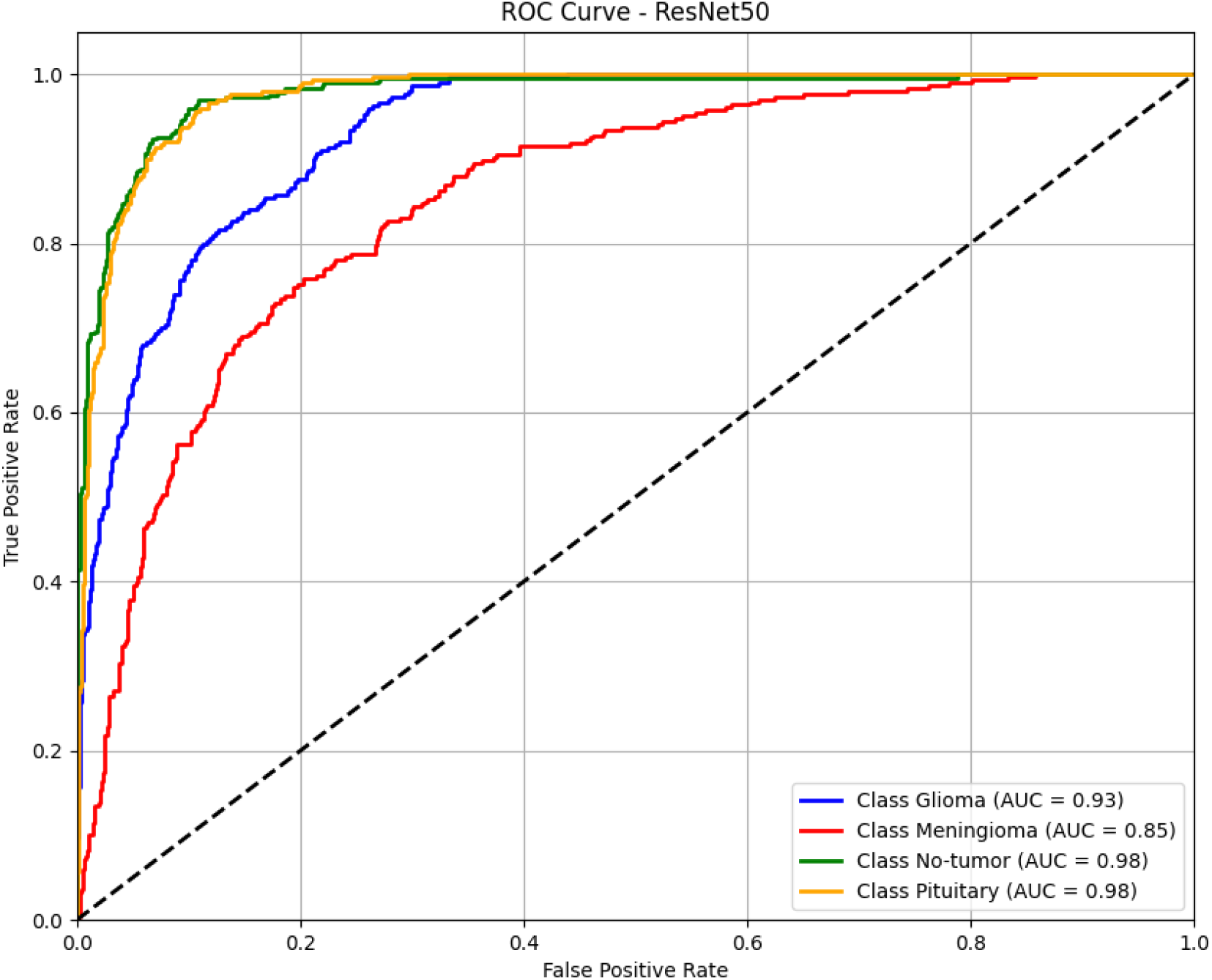

Figure 7 displays the ROC curve for DenseNet121. The model achieved perfect classification for all tumor types, with each class reaching an AUC of 1.00. This result indicates excellent sensitivity and specificity across all categories and highlights DenseNet121’s strong ability to differentiate between brain tumor types without misclassification [3,6].

**Figure 7.**
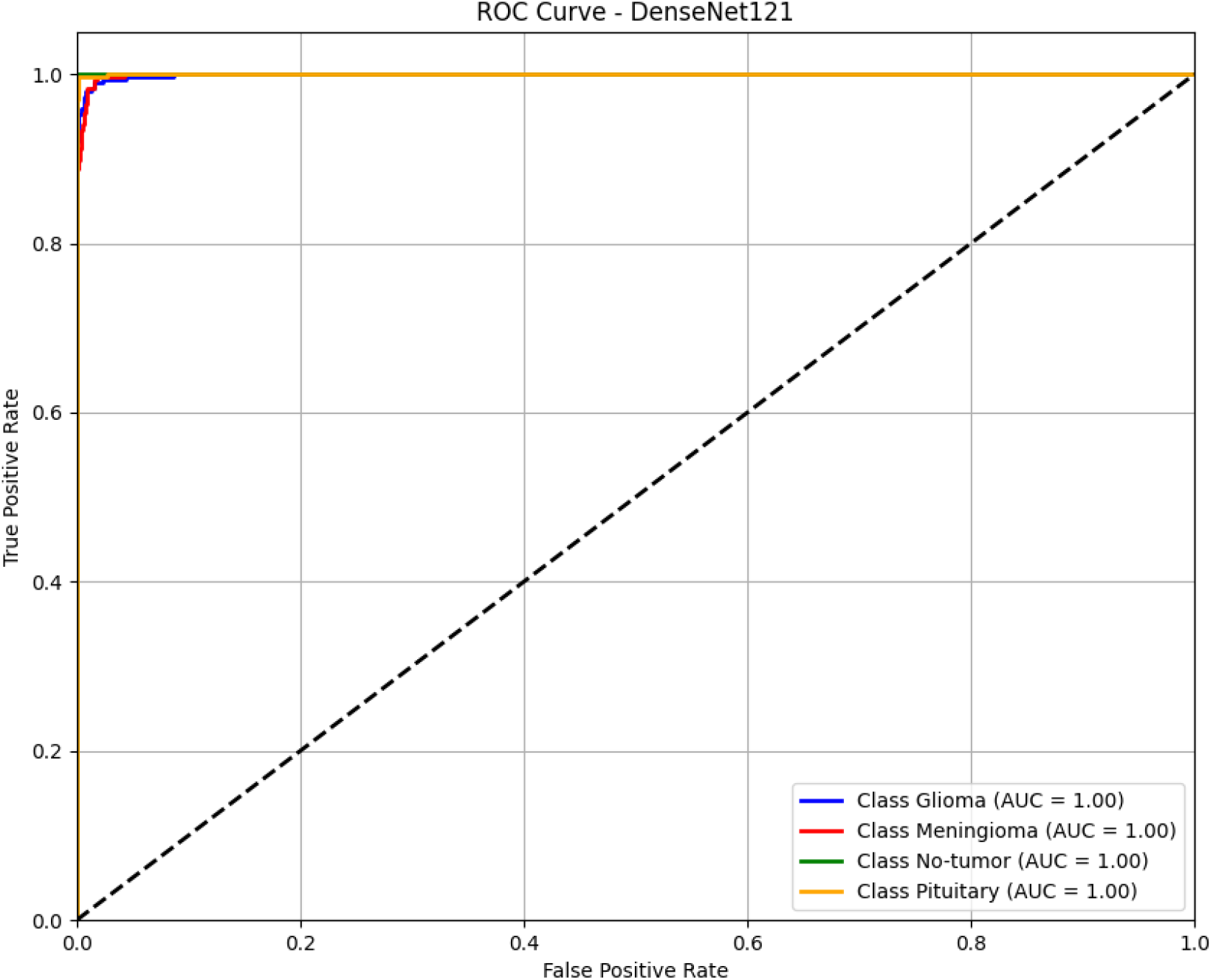

As shown in Figure 8, the XGBoost ensemble model achieved perfect classification results, with all four tumor classes reaching an AUC of 1.00. This confirms the model’s exceptional ability to distinguish between tumor types with no overlap or confusion, supporting its use in high-accuracy medical applications [4,6].

**Figure 8.**
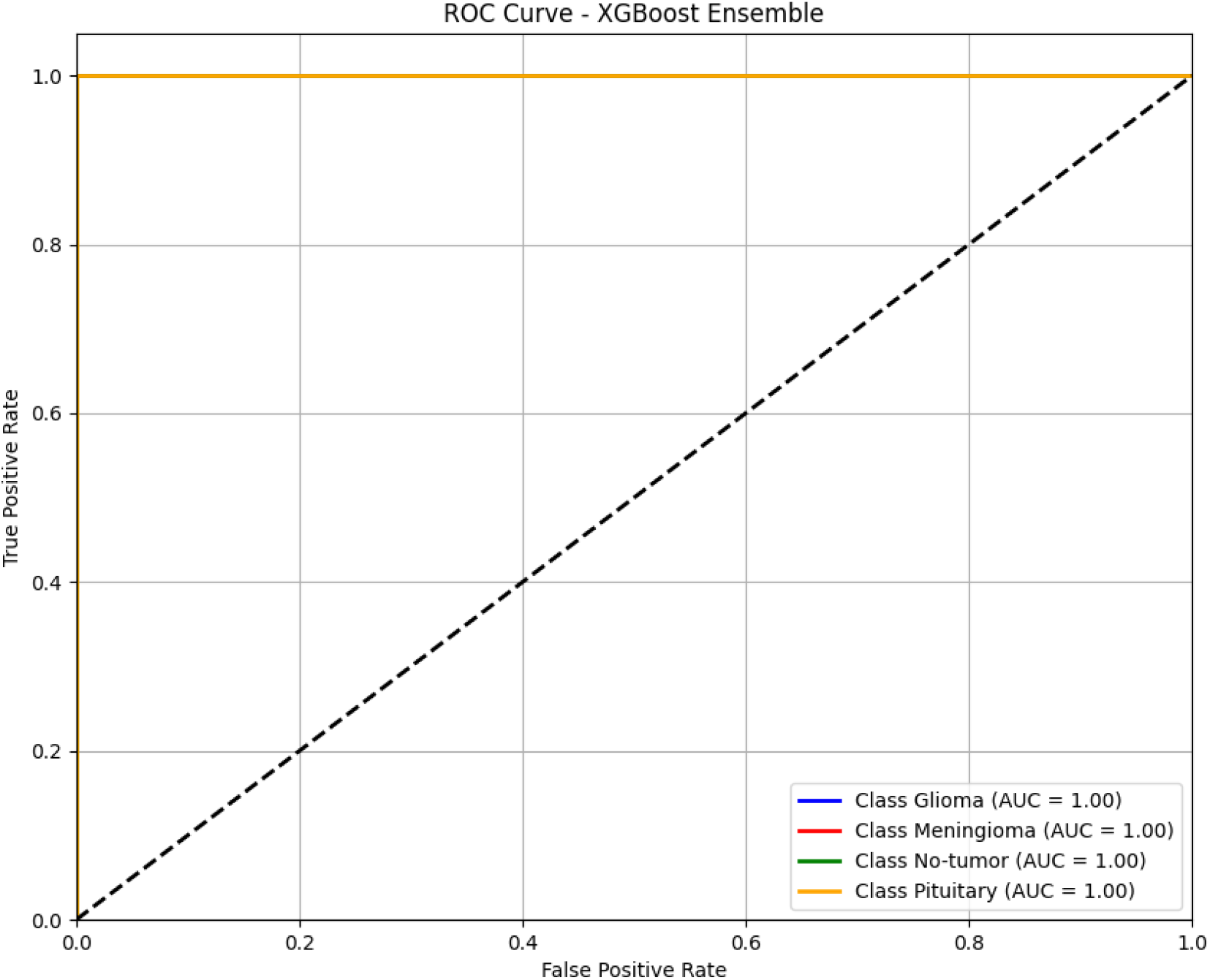

Figure 9 illustrates the ROC curve of the soft voting ensemble. Similar to the XGBoost model, all four tumor categories reached an AUC of 1.00, confirming perfect sensitivity and specificity. The consistent performance across all classes reinforces the robustness and reliability of ensemble learning via soft voting in tumor classification [5,6].

**Figure 9.**
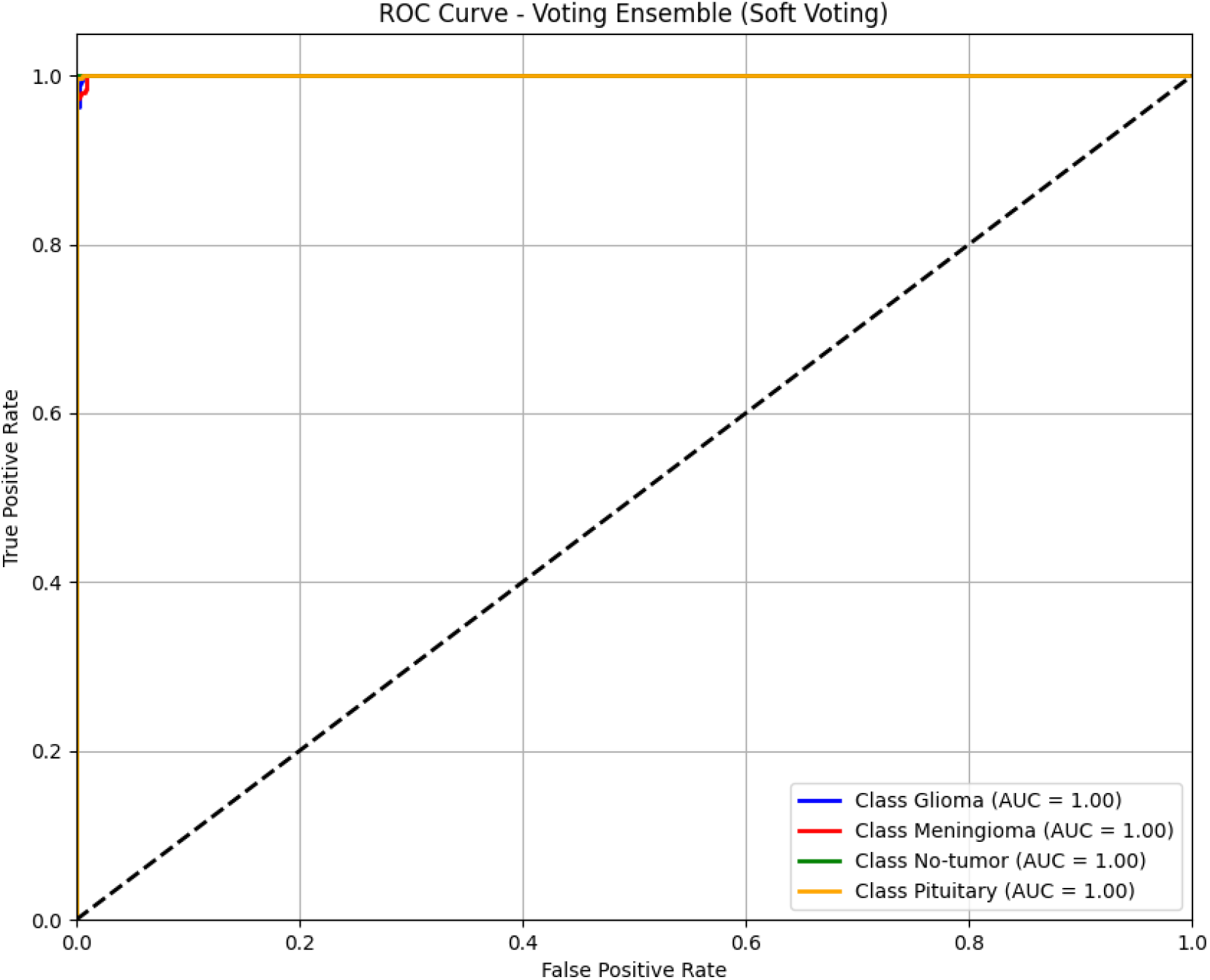

As shown in Figure 10, the stacking ensemble (with logistic regression as the meta-learner) achieved a perfect AUC of 1.00 for each class. This model demonstrates the strength of combining multiple base learners to produce a highly generalizable and accurate brain tumor classifier, suitable for deployment in clinical settings [5,6].

**Figure 10.**
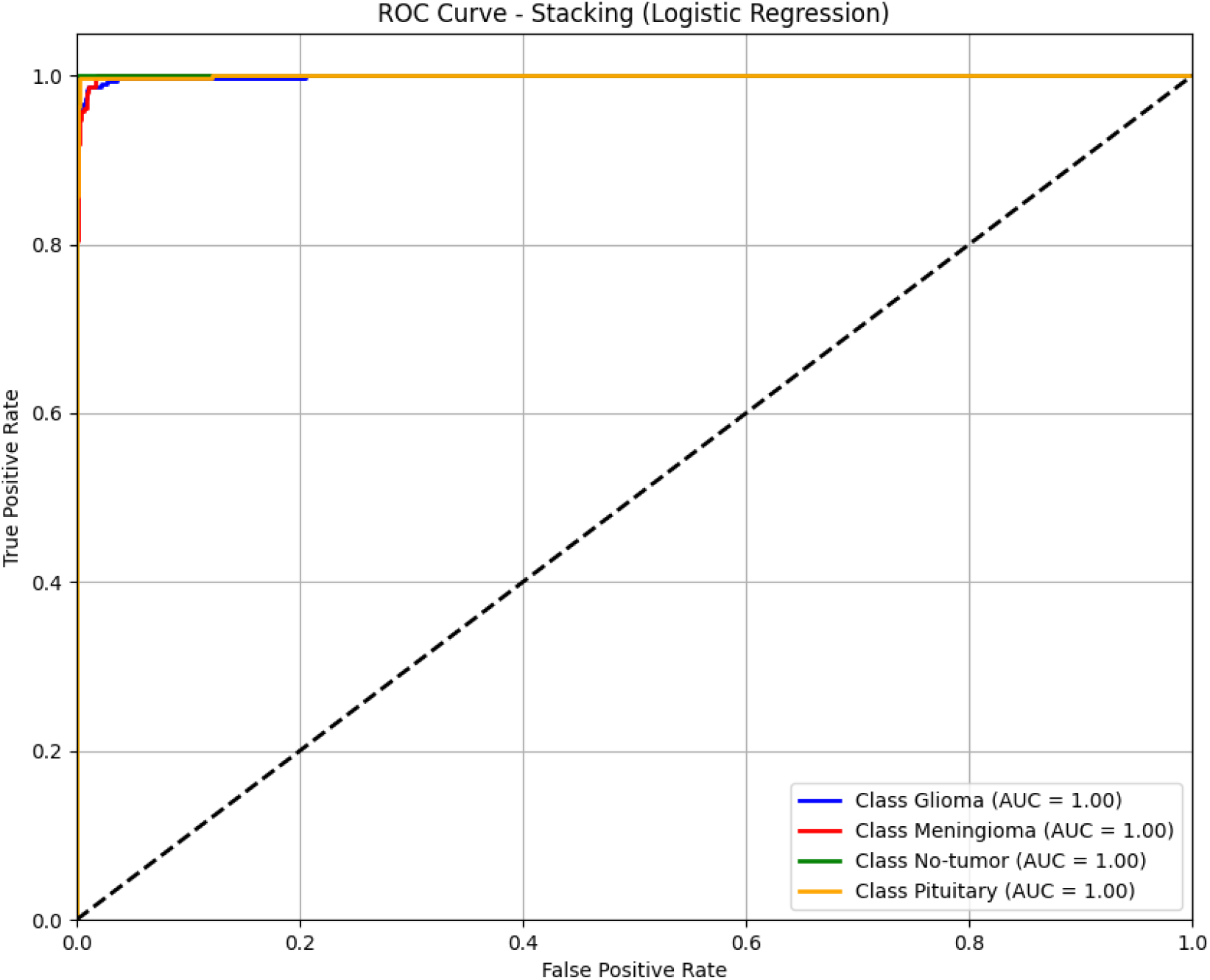

**Table 4.** Classification Report for Soft Voting Ensemble.

| Class | Precision | Recall | F1-Score | Support |
| --- | --- | --- | --- | --- |
| Glioma | 0.98 | 1.00 | 0.99 | 300 |
| Meningioma | 1.00 | 0.98 | 0.99 | 306 |
| No-tumor | 1.00 | 1.00 | 1.00 | 405 |
| Pituitary | 1.00 | 1.00 | 1.00 | 300 |

## Conclusion

In this study, we proposed and evaluated several deep learning-based models for multi-class brain tumor classification using MRI images. The fine-tuned VGG16 model achieved a high test accuracy of 99.24%, demonstrating strong performance across all tumor types [1]. Among the individual models, DenseNet121 and XGBoost achieved perfect classification performance, with all four tumor types (Glioma, Meningioma, No-Tumor, and Pituitary) classified with 100% accuracy [3,4]. Additionally, ensemble approaches such as soft voting and stacking further enhanced the robustness and generalization of the predictions, reaching overall accuracies of 99.54% and 98.47%, respectively [5]. The ROC curves and AUC values confirmed the effectiveness of the proposed methods in distinguishing between different tumor categories [6]. Overall, the results demonstrate that combining powerful CNN architectures with ensemble learning strategies can significantly improve the accuracy and reliability of brain tumor classification systems. These findings provide a strong foundation for developing effective computer-aided diagnosis tools in real clinical settings.

### Limitations and Future Work

Despite achieving high accuracy across models and ensemble methods, this study has some limitations.

First, the used dataset is relatively balanced and clean, which may not reflect the variability and noise encountered in real-world clinical data [11]. Moreover, the dataset consists of 2D MRI slices without specifying imaging planes, potentially limiting model generalization across different scan orientations [11]. Another limitation is that evaluation was conducted on a fixed internal dataset split and no external validation was performed, which may restrict the generalizability of the findings [6].

In future work, we aim to validate the proposed models on larger, more diverse, and multi-institutional datasets to ensure clinical applicability [11]. We also plan to evaluate models on multi-planar or 3D volumetric MRI data. Finally, integrating explainable AI techniques such as Grad-CAM or SHAP could provide deeper insights into the decision-making process of the models, enhancing their interpretability and trust in clinical settings [7].

## Data Availability

Data Availability Statement:
The data used in this study are publicly and freely accessible through the original source (link provided). These data are provided solely to verify the accuracy of the findings and to enable reproducibility of the results. Any unauthorized alteration, manipulation, or misuse of the data is strictly prohibited and subject to legal consequences.

https://www.kaggle.com/datasets/mohamadabouali1/mri-brain-tumor-dataset-4-class-7023-images/data

**Dataset**

MRI Brain Tumor Dataset 4-class (7023 images), Mohamad Abou Ali, Kaggle, 2021. [Online].

Available:

https://www.kaggle.com/datasets/mohamadabouali1/mri-brain-tumor-dataset-4-class-7023-images[11]

The experiments were conducted on the publicly available MRI Brain Tumor Dataset (4-class, 7023 images) provided by Mohamad Abou Ali on Kaggle [Online]. This dataset includes four tumor classes: Glioma, Meningioma, Pituitary tumor, and No tumor.

